# Structural and functional connectivity in infancy relate to communication skills at age 2 in children born very preterm

**DOI:** 10.64898/2026.06.13.26355553

**Authors:** Jennifer Vannest, Mekibib Altaye, Junqi Wang, Maria E. Barnes-Davis, Lili He, Nehal A. Parikh, Lisa Hunter

## Abstract

Preterm birth is associated with increased risk for communication difficulties, yet early neural markers of later outcomes remain poorly understood. This study examined associations between structural and functional brain connectivity in infancy and communication skills at 24to 30 months in children born very preterm (≤32 weeks gestation). Participants (n = 180) MRI at term-equivalent age during natural sleep. Resting-state functional and diffusion data were used to derive structural and functional connectivity across regions implicated in communication. At follow-up, communication outcomes were assessed using Communication and Symbolic Behavior Scales (CSBS), which captures verbal, gestural, social-affective, and symbolic communication skills.

We analyzed 22×22 functional and structural connectomes among selected ROIs, using a LASSO regression approach to identify connectivity associated with CSBS scores adjusting for demographic and medical covariates. Significant relationships were observed between both functional and structural connectivity and communication skills, differing by domain. Functional connectivity between left and right temporal regions was positively associated with overall communication scores, whereas several structural connections involving cortical, cerebellar, and subcortical regions showed negative associations. Distinct connectivity patterns were also associated with gestural, verbal, social-affective, and symbolic communication skills.

These findings demonstrate that variability in early brain connectivity is associated with later communication outcomes in children born very preterm. We found both significant positive and negative associations after adjusting for relevant covariates; these patterns potentially reflect compensatory or atypical network organization. These results highlight the value of multimodal neuroimaging in identifying early neural correlates of communication in this high-risk population.

**Highlights:** Early structural and functional brain connectivity is associated with communication outcomes at 2–3 years in children born very preterm.

Distinct patterns of connectivity relate differently to verbal, gestural, social-affective, and symbolic communication skills.

Both positive and negative connectivity–behavior relationships suggest complex and potentially compensatory neural developmental pathways.

Multimodal neuroimaging may provide early biomarkers to support identification and intervention for communication risk in preterm populations.

## Introduction

Preterm birth is a significant risk factor for adverse developmental outcomes, particularly in the domain of communication skills. More than 450,000 preterm babies survive every year in the US: 11% of all births [1, 2]. Preterm infants have a 3 to 10-fold increase in speech-language and hearing deficits compared to children born at full-term. About 40% of very preterm infants (≤32 weeks gestational age) develop persistent speech or language difficulties (SLD) while 4-10% have permanent hearing loss, increasing the impact on communication [3–6]. Currently, it is not possible to accurately predict in the first year of life which children will develop SLD. As a result, speech-language therapy is the most delayed and the least common of the therapy services provided to infants after NICU discharge. In addition, preterm children may undergo limited follow-up assessment of communication skills in the first two years of life. Children that are assessed typically complete the Bayley Scales of Infant and Toddler Development, the current clinical standard for guiding early intervention in preterm infants. However, there is concern that this tool overestimates cognition and language and is not predictive of later impairment in term or preterm children [7].

Neuroimaging studies have begun to elucidate the neural basis of SLD in children born preterm. At term-equivalent age (TEA), infants born preterm have been shown to have leftward asymmetries in volume and diffusion in parts of the superior longitudinal fasciculus and corticospinal tracts [8–10], similar to term-born children. However, while these anatomical asymmetries may not be grossly affected by preterm birth, differences in both structural and functional connectivity between preterm and term-born infants have been observed [11–18]. In term-born infants, bilateral but left-lateralized networks connecting frontal and temporal regions have been established [16, 19–21], forming the initial architecture for dorsal and ventral language pathways present in adults [22–25]. Studies comparing preterm and term infants suggest complex differences in brain network organization and altered patterns of development, including a lesser degree of left-lateralization and reduced interhemispheric connectivity, which may underlie differences in language development. Similarly, in older children and adults with a history of preterm birth, patterns of both increased and decreased connectivity relative to term-born controls have been observed [13, 26–29], Interhemispheric and cerebellar-cortical functional connections have been shown to relate to better language skills, and structural connectivity results suggest an increased reliance on extracallosal pathways in childhood. [30, 31]

However, there is minimal longitudinal data available to understand how functional and structural connectivity in preterm infants provide an early marker of communication skills in childhood. In term-born children, connectivity patterns of left temporal regions in infancy have been shown to relate to language skills at school age [32], but this relationship is likely to be altered in preterm infants. In a large sample of preterm infants, Barnes-Davis et al. related structural connectivity at TEA to language scores on the Bayley at age 2 and found negative relationships with connectivity in bilateral cerebellar white matter and middle cerebellar peduncles, bilateral corticospinal tracks, posterior commissure and the posterior inferior fronto-occipital fasciculus [26]. Most recently, Canini et al. [33] related functional connectivity in preterm infants to scores on the Bayley at 6, 12, 24, and 36 months. They found that functional connectivity between the cerebellar hemispheres predicted social-emotional scores on the Bayley at 6 months; connectivity between regions related to language and emotional control predicted social-emotional scores at 12 months; and connectivity between sensorimotor and higher-order control regions predicted Bayley language scores at 24 months and cognitive scores at 36 months. In line with the structural connectivity results reported above, they found a *negative* relationship between connectivity between auditory and language regions and social-emotional and cognitive scores at 36 months.

Overall, the literature suggests that preterm birth may result in patterns of connectivity in infancy that relate to variability in communication outcomes in early childhood, both in comparison to term children and within the preterm population. However, this relationship has not been assessed in very preterm children for whom both early life structural and functional connectivity and later communication skills have been assessed in a comprehensive, detailed way. In the present study, we examined structural and functional connectivity at TEA among brain regions known to underlie communication in a large sample of very preterm infants.

Importantly, we used the Communication and Symbolic Behavior Scales (CSBS) to assess a wide range of communication skills at 24-30 months corrected age. The CSBS is a direct, standardized assessment particularly well-suited to this population as it assesses gestures and other non-verbal communication, therefore allowing for a range of performance in children who are delayed in using words. The CSBS has good psychometric properties and has been shown in 2-yr-olds to predict expressive vocabulary one year later [34]. We also completed comprehensive hearing assessment on the same children at the same age as the language outcomes were measured.

We hypothesized that there would be associations between brain connectivity in preterm infants at term-equivalent age and later communication skills, after accounting for demographic and medical factors. Identifying these associations represents a step forward in understanding the underlying neural bases of effects of preterm birth on communication skills.

## Methods

### Participants

Participants were recruited from five Level 3 or 4 NICUs in the Greater Cincinnati area. Inclusion criteria included gestational age ≤32 weeks and written informed consent of parents/guardians. Exclusion criteria included known cyanotic congenital heart disease and genetic syndromes or congenital anomalies known to impact the CNS.

### Participant medical history and demographics

Demographic and medical variables including post-menstrual age (PMA) at time of MRI scan, gestational age and history of bronchopulmonary dysplasia, child race, and child sex were documented via medical chart review, and maternal education was assessed via parent report.

### Neuroimaging Protocol

All MRI scans were completed at 41-44.9 weeks PMA, during natural sleep after swaddling and feeding 30 minutes prior to the scan (no sedation was used). Silicone ear plugs were used for hearing protection. T1-weighted anatomical MRI was obtained using 3D FFE, TR/TE/TI 7.3/3.4/1610 ms, and resolution 1×1×1 mm. High-resolution T2-weighted MRI was obtained with TR/TE = 19 377/166 ms and resolution 1×1×1 mm. Diffusion MRI was obtained at b=0, TR/TE 6755/88 ms; b=800 s/mm2, TR/TE 6972/88ms, 36 directions; and b=2000 s/mm2, TR/TE =5073/88 ms, 68 directions, multi-band acceleration factor 4; all resolution 2×2×2 mm. Resting-state functional MRI (rsfMRI) was acquired with echo-planar imaging, TR/TE 1187/23 ms, resolution 2.5×2.5×2.5 mm, 400 frames, multi-band acceleration factor 3.

### Communication assessment at 24-30 months corrected age

All children were administered the Communication and Symbolic Behavior Scales (CSBS) to assess communication skills. Specifically, administration of the CSBS is divided into three main sections: first, a series of communication temptations, which include toys and books that prompt children to request help from adults (Scales 1–18). The second section involves pretend play to assess play schemes and comprehension (Scales 19–21); and **t**he final section involves constructive play (Scale 22). Each scale is scored from 1 to 5, and scales are summed to create cluster scores.

The first cluster score summarizes Communicative Functions, including behavior regulation (Scale 1), where the child requests an adult to perform an action; joint attention (Scale 2), where the child directs the adult’s attention to an object or event; and sociability (Scale 3), which measures the percentage of communicative acts that are social rather than joint attention. The Gestural Communication cluster score reflects children’s use of conventional gestures like pointing and nodding (Scale 4), distal gestures toward out-of-reach objects (Scale 5), and gestures combined with vocalizations (Scale 6). The Vocal Communication cluster score is tabulated based on vocal acts not aligned with gestures (Scale 7), consonant inventory (Scale 8), the number of syllables with consonants (Scale 9), and multisyllabic utterances (Scale 10). The Verbal Communication cluster score most resembles traditional tests of language development as it is based on the number of words expressed (Scale 11) and number of word combinations (Scale 12).

The Reciprocity cluster assesses communication with the adult and includes communication in response to the adult (Scale 13), the child’s rate of communicative acts (Scale 14), and repair strategies (Scale 15), which involve repeating or modifying communication during temptations. The Social-Affective Signaling cluster score is measured through the child’s gaze shifts between the adult and play objects (Scale 16), shared positive affect (Scale 17), and negative affect (Scale 18). Finally, the Symbolic Behavior cluster score is based on assessing the child’s language comprehension during play (Scale 19), an inventory of the child’s play action schemes (Scale 20), complexity of these schemes (Scale 21), and constructive play (Scale 22). The Communication Composite Score is calculated by summing the scaled scores from Scales 1 through 18.

Although CSBS is standardized for children up to 24 months, it can also be used for older children with developmental delays. Because children in our study were 24-30 months corrected age, we used raw cluster scores and included the child’s corrected age at testing in our analyses.

The CSBS takes approximately 40 minutes to administer in a session with the parent/caregiver and child. The CSBS was video recorded and later scored by trained study staff. Double scoring for 20% of assessments scored by video was completed for inter-rater reliability; total scores had good reliability (ICC=0.77).

The Bayley Scales of Infant and Toddler Development (Version III or IV) were also administered to a subset of participants that were available to participate in an additional testing session at 22 to 26 months corrected age.

### Hearing assessment at 24-30 months corrected age

Hearing was assessed using both behavioral and physiologic hearing tests to classify type and severity of hearing loss, as described previously [35]. The behavioral tests included speech detection threshold for two-part spoken words and hearing threshold for pure tone stimuli at 1, 2, 4 and 8 kHz. The technique used was conditioned play audiometry if the child was developmentally able to follow directions, or visual reinforced audiometry if not. Insert earphones were used to obtain thresholds in both ears, unless the child would not tolerate them, in which case stimuli were presented to both ears using speakers. Physiologic tests included tympanometry to assess middle ear function, and otoacoustic emissions to assess inner ear function. Speech reception thresholds and pure tone thresholds averaged for both ears and across all frequencies obtained were used as the primary hearing variables in this analysis.

## Data Analysis

Analyses focused on the CSBS Communication Composite raw score and selected raw cluster scores on the CSBS; specifically, Gestural Communication, Verbal Communication, Social-Affective Signaling, and Symbolic Behavior. We selected these clusters in order to investigate the neural underpinnings of skills included on typical language assessments in this age range (Verbal Communication), but also to uniquely examine those skills frequently excluded from early communication assessments (Gesture, Social-Affective, and Play). For those participants for whom Bayley scores were available, we used Pearson correlation to examine the relationship between these four CSBS cluster scores of interest and Bayley Cognitive and Language composite scores (Bonferroni corrected to p<.006).

Neuroimaging data were preprocessed using tools from the FMRIB Software Library (FSL) [36] and the developing Human Connectome Project (dHCP). Out of the 82 default regions in the dHCP atlas [37], we selected 22 predefined regions of interest (ROIs) for analysis, including frontal, temporal, and parietal cortices, along with subcortical regions and cerebellum, all implicated in communication. Regions of interest are listed in Table 1. Diffusion MRI (dMRI) data were processed using FSL to correct for off-resonance field effects, eddy current distortions, and motion [38, 39]. Fractional anisotropy (FA) was calculated [40] with reconstruction of fiber pathways [41]. To derive each infant’s structural connectome, we computed the mean functional anisotropy values for all fiber tracts connecting 22×22 selected ROIs. To derive each infant’s structural connectome, probabilistic tractography was initiated from each of the 22 selected seed ROIs. Tracking was anatomically constrained to each infant’s reconstructed white matter tissue volume. Structural connections were defined as reconstructed white matter pathways connecting pairs of selected ROIs. For each successfully reconstructed ROI-to-ROI pathway, we computed the mean fractional anisotropy, resulting in a 22 × 22 structural connectivity matrix for each participant.

**Table 1.**
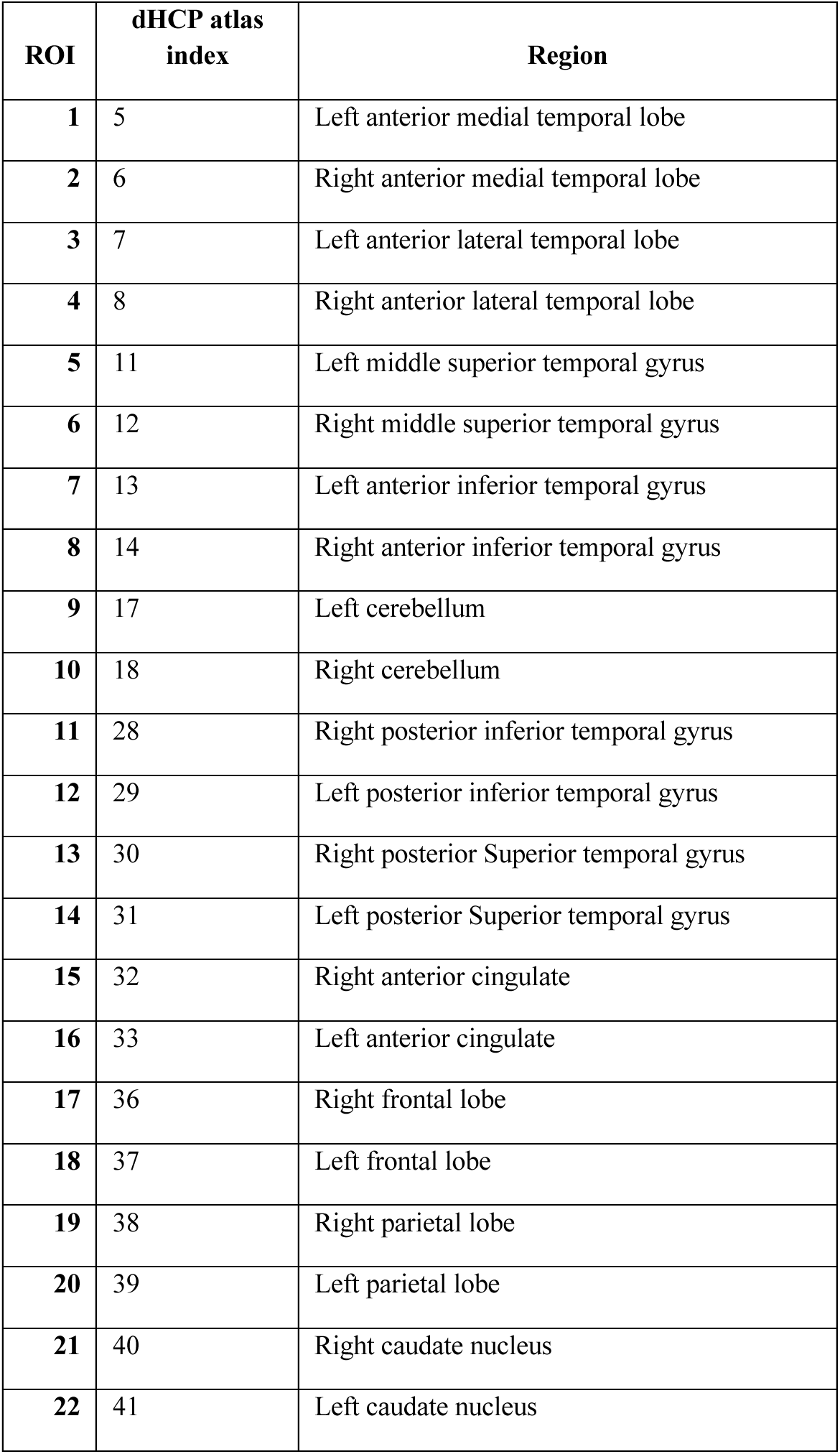
Regions of interest for connectivity analyses.

Resting-state fMRI data were preprocessed using the dHCP functional pipeline [42] including distortion correction, slice-to-volume motion correction, and structured denoising. Pearson correlations between each pair of ROIs were computed, generating a 22×22 functional connectome.

Pairwise functional and structural connections were used as input to a penalized regression approach using the Least Absolute Shrinkage and Selection Operator (LASSO). The goal of this analysis was to identify a stable and generalizable set of functional and structural connections associated with communication outcomes measured by the CSBS. To evaluate the robustness of variable selection and variability in model performance, we generated 500 bootstrap samples by resampling participants with replacement from the original dataset. Within each bootstrap iteration, a LASSO regression model was fitted using 5-fold cross-validation to select the regularization parameter. Candidate connections were retained for second-stage analysis if they were selected in at least 80% of the 500 bootstrap iterations.

In a second-stage analysis, candidate connections identified from the bootstrap LASSO procedure were evaluated in a multivariable regression framework to determine whether they remained associated with CSBS outcomes after adjustment for prespecified demographic and medical covariates. Six a-priori covariates were included in all models: maternal education, child sex, postmenstrual age at MRI scan, corrected age at CSBS testing, bronchopulmonary dysplasia severity (0-3 scale, ranging from none to severe [43]), and Global Brain Abnormality Score as described by Kidokoro et al. [44]. Maternal education was modeled as a three-level categorical variable: high school or lower, college, and graduate education. Candidate functional and structural connections selected from the LASSO stability procedure were then entered as additional predictors and allowed to enter or leave the model through stepwise selection. This approach was used to identify which connections remained associated with each outcome after adjustment for the prespecified demographic and medical variables and to quantify their additional contribution to outcome variability. Associations retained in the final stepwise regression model were considered statistically significant at p < .05.

## Results

180 children completed both neuroimaging at TEA and communication assessment using the CSBS at age 24-30 months corrected age. Demographic, hearing and medical variables are summarized in Table 2. 144 children completed the Bayley Cognitive assessment and 143 completed the Bayley Language assessment. Mean CSBS raw scores and Bayley standard scores are included in Table 3. 155 participants had functional imaging data that was of sufficient quality for analysis, and 159 participants had structural imaging data that was of sufficient quality for analysis.

**Table 2.** Demographics, medical and hearing factors for all participants. n=180 except where noted.

|  |  |
| --- | --- |
| Sex (M/F) | 92/88 |
| Race (n per caregiver report) | Black/African American =34; White=131; Multiple Races=11; not reported=4 |
| Maternal education (n per caregiver report) | High school=35; College=92; Graduate=40; not reported=13 |
| Gestational age (weeks; mean, SD) | 29.76 (2.30) |
| Postmenstrual age at MRI (weeks; mean, SD) | 42.77 (1.52) |
| Global Brain Abnormality Score [44] (mean, SD) | 5.18 (3.49) |
| BPD grade (n per category) | none=112; mild =34; moderate=34 |
| SAT both ears (mean, SD), n=175 | 10.53 dB HL* (8.35) |
| PTA both ears (mean, SD), n=176 | 14.44 dB HL* (9.42) |

- dB HL = decibels hearing level, referenced to 0 dB HL as normal average hearing level in young adults.

**Table 3.** CSBS and Bayley scores; n=180 except where noted.

|  |  |
| --- | --- |
| CSBS Gestural Communication raw cluster score (max 15; mean, SD) | 9.54 (2.74) |
| CSBS Verbal Communication raw cluster score (max 10; mean, SD) | 7.73 (1.35) |
| CSBS Social Affective raw cluster score (max 15; mean, SD) | 9.82 (1.72) |
| CSBS Symbolic Behavior raw cluster score (max 20; mean, SD) | 15.42 (2.62) |
| CSBS total raw score (max 90; mean, SD) | 61.29 (10.18) |
| Bayley Cognitive Composite standard score (n=144, mean, SD) | 94.15 (16.08) |
| Bayley Language Composite standard score (n=143, mean, SD) | 94.28 (18.27) |

### Relationships between CSBS raw cluster scores and Bayley Composite Standard Scores

The CSBS Verbal Communication raw cluster score correlated significantly with Bayley Cognitive scores (r=0.22, p=.006) and also showed a stronger correlation with Bayley Language scores (r=0.41, p<.001). The CSBS Social-Affective Signaling raw cluster score also correlated positively with Bayley Language scores (r=.26, p=.002). The Symbolic Behavior raw cluster score correlated positively with Bayley Cognitive scores (r=0.41, p=.006) and with Bayley Language scores (r=0.34, p<.001).

### Relationships between imaging at TEA and CSBS raw scores at age 2

#### CSBS Communication Composite Raw Score

Significant relationships were observed between functional and structural connectivity and CSBS Communication Composite raw scores. First, functional connectivity between the right middle superior temporal and left anterior inferior temporal region was positively related to Communication Composite raw scores. In contrast, functional connectivity between the right frontal lobe and left anterior inferior temporal region, structural connectivity between the right cerebellum and left posterior superior temporal gyrus, as well as structural connectivity between the right frontal lobe and right caudate nucleus related negatively to Communication Composite raw scores.

#### CSBS Gestural Communication

Significant relationships were observed between maternal education and Gestural Communication raw cluster scores. Structural connectivity between the right anterior medial temporal lobe and left anterior lateral temporal lobe, and the right middle superior temporal and right posterior inferior temporal cortices showed a positive relationship with Gestural Communication raw cluster scores, and structural connectivity between the right middle superior temporal cortex and left caudate was negatively related to Gestural Communication cluster scores.

#### CSBS Verbal Communication

A significant positive relationship was observed between age at testing and CSBS Verbal Communication cluster scores. Structural connectivity between the left cerebellum and left posterior inferior temporal cortex was positively related to CSBS Verbal Communication cluster scores. Significant negative relationships with CSBS Verbal Communication cluster scores were observed for structural connectivity between the left anterior inferior temporal and left posterior inferior temporal regions, and between right anterior inferior temporal and right frontal cortices.

#### CSBS Social-Affective Signaling

Functional connectivity between the left frontal lobe and left parietal lobe was positively related to CSBS Social-Affective Signaling cluster scores. Significant negative relationships with CSBS Social-Affective Signaling cluster scores were observed for structural connectivity between left frontal lobe and right caudate nucleus.

#### CSBS Symbolic Behavior

Significant relationships were observed between BPD grade and child sex, and CSBS Symbolic Behavior cluster scores. Functional connectivity between the right anterior inferior temporal and right frontal cortices was positively related to CSBS Symbolic Behavior cluster scores, and functional connectivity between the right and left parietal cortices was negatively related to CSBS Symbolic Behavior cluster scores.

A significant positive relationship with CSBS Symbolic Behavior cluster scores was observed for structural connectivity between the left medial anterior temporal region and right caudate nucleus, and left posterior inferior temporal gyrus and left posterior superior temporal gyrus. Negative relationships with CSBS Symbolic Behavior cluster scores were found for three structural connections: between the right medial anterior temporal cortex and right anterior cingulate, between right cerebellum and right frontal cortex, and between the left medial anterior temporal and right anterior inferior temporal cortices.

## Discussion

These results demonstrate that there is an underlying neural basis supporting a wide range of communication skills in very preterm children, detectable in patterns of functional and structural connectivity at TEA. Interestingly, consistent with other work in this area, there were a number of positive and negative relationships between structural and functional connectivity at TEA and communication outcomes at age 2. Assessment using the CSBS allowed us to capture children’s skills in verbal expression as well as gesture, social-affective signaling, and symbolic play, and assess patterns of connectivity in infancy associated with each of these subscales.

Communication composite scores on the CSBS were positively associated with increased functional connectivity between regions of the left and right temporal lobes. However, structural connectivity between the same region of the anterior inferior temporal lobe and the right frontal lobe was negatively associated with composite scores, as well as a connection between the right frontal lobe and caudate nucleus. Structural connectivity between the left posterior temporal lobe and right cerebellum was also negatively correlated. Because the CSBS composite score is composed of subscales assessing varied aspects of communication that may have a varied neural basis, examining relationships of connectivity with subscale scores may allow for clearer interpretation.

Specifically, gestural communication skills were associated with connectivity between bilateral temporal regions, and with greater connectivity between regions of the right temporal lobe. While this specific relationship has not previously been studied in infants or young children, studies of adults have shown bilateral temporal engagement in gesture comprehension [45–48]. In contrast, an interhemispheric, cortico-striatal connection was negatively related to gestural communication.

For verbal skills, a left temporal – left cerebellar structural connection was positively associated with these scores; previous studies have noted the reliance on cerebellar pathways in relation to language skills in children with a history of preterm birth [27, 28, 49]. However, within-hemisphere connections in both hemispheres were negatively associated with later verbal outcomes, consistent with earlier findings relating connectivity in infancy to later language scores [26, 33]; these connections involved inferior temporal regions that, in adults, support visual and semantic processing.

In contrast, social-affective signaling was positively related to left fronto-parietal functional connectivity. Social aspects of communication are complex and rely on mechanisms that develop rapidly in the first years of life [50]. One neuroimaging study of 12- and 24-month olds found that dorsal attention network connectivity was related to joint attention, one component of social communication [51]; another study of preschoolers found that fronto-parietal connectivity was reduced in children with autism and attention difficulties [52]. As with gestural communication, an interhemispheric, cortico-striatal connection was negatively related to social-affective signaling.

The pattern of results relating connectivity in infancy to symbolic behavior differed in some ways from other skills, but similar to gestural communication, symbolic behavior was positively associated with right hemisphere intra-hemispheric functional connectivity between right anterior-inferior temporal and right frontal cortices, however, a structural right hemisphere connection between right medial anterior temporal cortex and right anterior cingulate was negatively related. In contrast to gesture and social-affective skills, an interhemispheric, cortico-striatal structural connection was positively related to symbolic play. Other interhemispheric connections between regions of cortex, as well as structural connectivity between right cerebellum and right frontal cortex were negatively related to symbolic behavior.

Broadly, these results support the hypothesis that variability in structural and functional connectivity underlies the variability in communication skills observed in children with a history of very preterm birth. Adult-like functional networks are just beginning to develop in early infancy; primary sensorimotor, auditory and visual networks have been shown to be most well-established in infants [53, 54]. Our focused set of ROIs related to communication included regions of the auditory network – connections involving right auditory cortex with other cortical regions in infancy were positively related to communication skills at age 2. Higher-order networks supporting default-mode, salience and attention have also been observed in infants, though these networks may be more variable in infancy ([55–57], see Carnevali et al. [58] for a recent review of functional imaging studies). Some of our results reflect a foundation for communication skills as these higher-order networks are established (e.g. increased left fronto-parietal functional connectivity associated with improved social communication). However, preterm birth has been shown to broadly affect structural and functional network architecture [10, 14–18, 26]. Negative associations we uncovered between communication skills and connectivity between cortical regions that are not typically associated with dorsal or ventral language pathways [25], nor with canonical higher-order networks, may reflect atypical pathways in infants born preterm.

Subcortical regions have also been suggested to act as important hubs in network organization in the neonatal period [55, 57], and their role has been suggested to shift during the first years of life [59], which may explain why we found variable relationships between cortical-caudate connectivity in infancy and later communication skills. In older children, abnormalities in basal ganglia structure and function have been suggested to underlie developmental language disorder [60].

In addition, the cerebellum has been suggested to be an important compensatory pathway supporting language skills in older children and young adults with a history of preterm birth [27, 28], and our results showing a positive relationship between verbal subscale scores and left cerebellum-inferior temporal structural connectivity support this hypothesis. However, structural connectivity of the right cerebellum with other regions of cortex was negatively associated with communication composite scores and symbolic behavior; this is consistent with similar work in preterm infants showing negative relationships between cerebellar structural connectivity and language outcomes on the Bayley-III at age 2 [26]. Barnes-Davis et al. suggest that increased cerebellar connectivity may represent a supplemental language pathway that may be established in some preterm infants earlier in development, then by school-age is supportive of better language skills. Thus, the timing of increased connectivity in cerebellar white matter could have developmental implications, with earlier reliance on this corticocerebellar network being deleterious to language development long-term. It should be noted that those previously published studies were in extremely preterm children and did not include neuroimaging from infancy and from childhood for the same participants [26, 28]. Future work should include neuroimaging from infancy and childhood within the same individuals to assess true developmental trajectories, as well as to better understand differences between right and left cerebellar connectivity and how different aspects of communication are supported by cortico-cerebellar connectivity.

The present analysis focused on a region-of-interest-based connectivity approach to examine connectivity patterns within putative networks supporting a range of foundational communication as assessed by the CSBS at age 2. This approach allowed us to examine pairwise connectivity among these crucial brain regions, however it did not consider connectivity through the whole brain. An additional limitation is that children’s communication skills, while assessed comprehensively by the CSBS, may still be highly variable at age 2 years. Longitudinal assessment at ages 3 years and above would better characterize whether children with a history of preterm birth have communication delays or difficulties. Our multimodal neuroimaging was obtained at TEA and was not repeated in the toddler or preschool age range. Future work should consider longitudinal assessment of brain connectivity. Additionally, our functional neuroimaging at TEA was obtained during resting state only, not during language tasks. Nonetheless, we feel the results we report are meaningful.

In conclusion, we report complex associations between functional and structural connectivity in the brain at term equivalent age and language and communication skills at 2-3 years of age in a prospective birth cohort of children born very and extremely preterm. Our study is unique in the inclusion of connectivity metrics from both structural and functional MRI, in the inclusion of not only very preterm but also extremely preterm and periviable children with gestational ages at birth as early as 23 weeks, in the addition of not only early hearing and audiological assessments in infancy but also during the toddler years, and in the inclusion of medical and socioenvironmental variables which are known to impact language attainments in children born preterm. This cohort includes children with and without a history of late talking and/or SLD. Future work will follow these children to determine the natural history of brain and language development in preterm children with SLD. Understanding the neural bases of these delays or difficulties may allow for earlier, targeted intervention and promote improved neurodevelopment in children following preterm birth.

**Figure 1.**
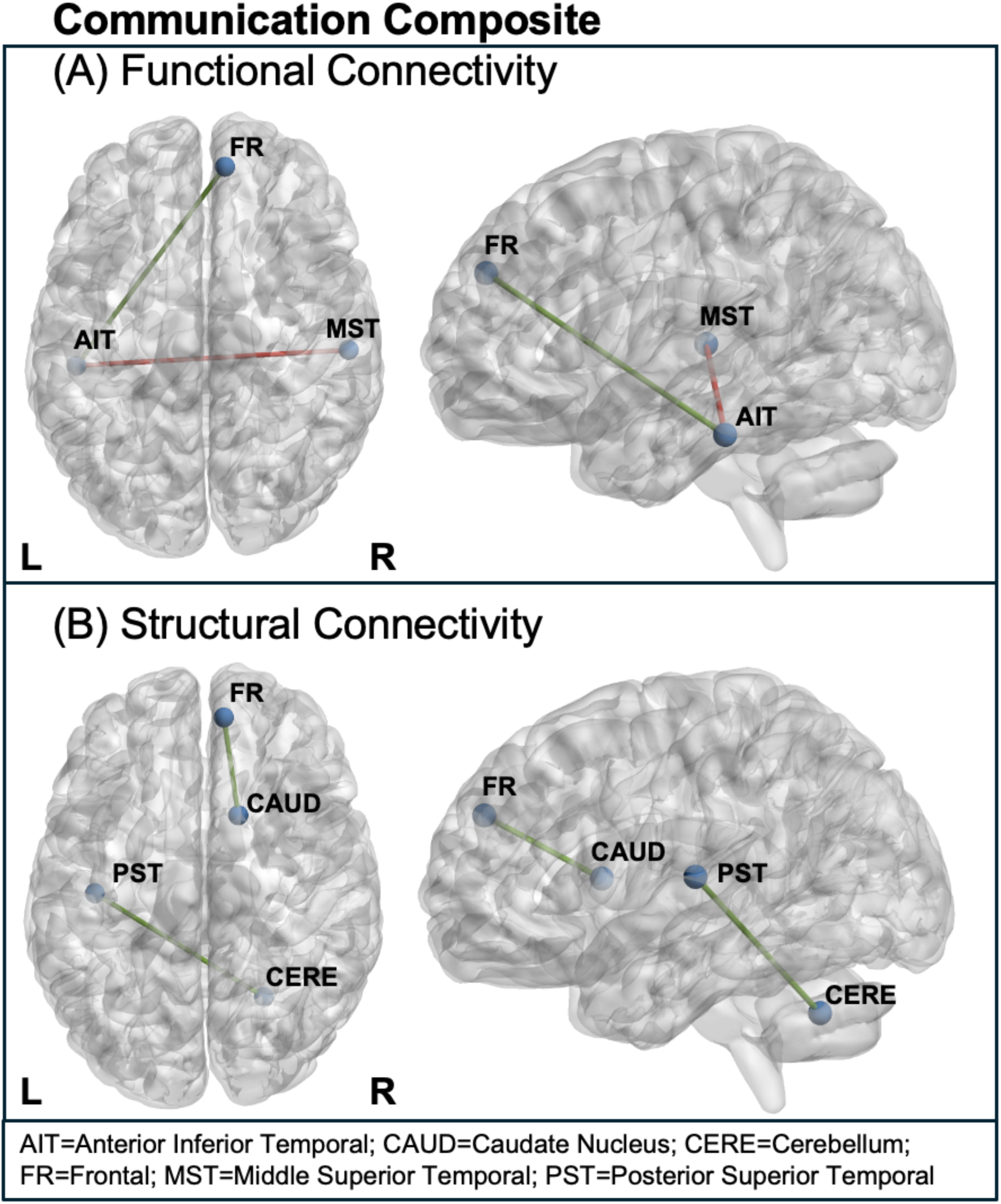
Pairwise (A) functional and (B) structural connections at TEA associated with Communication Composite raw scores at age 2. Red = positive associations, Green = negative associations

**Figure 2.**
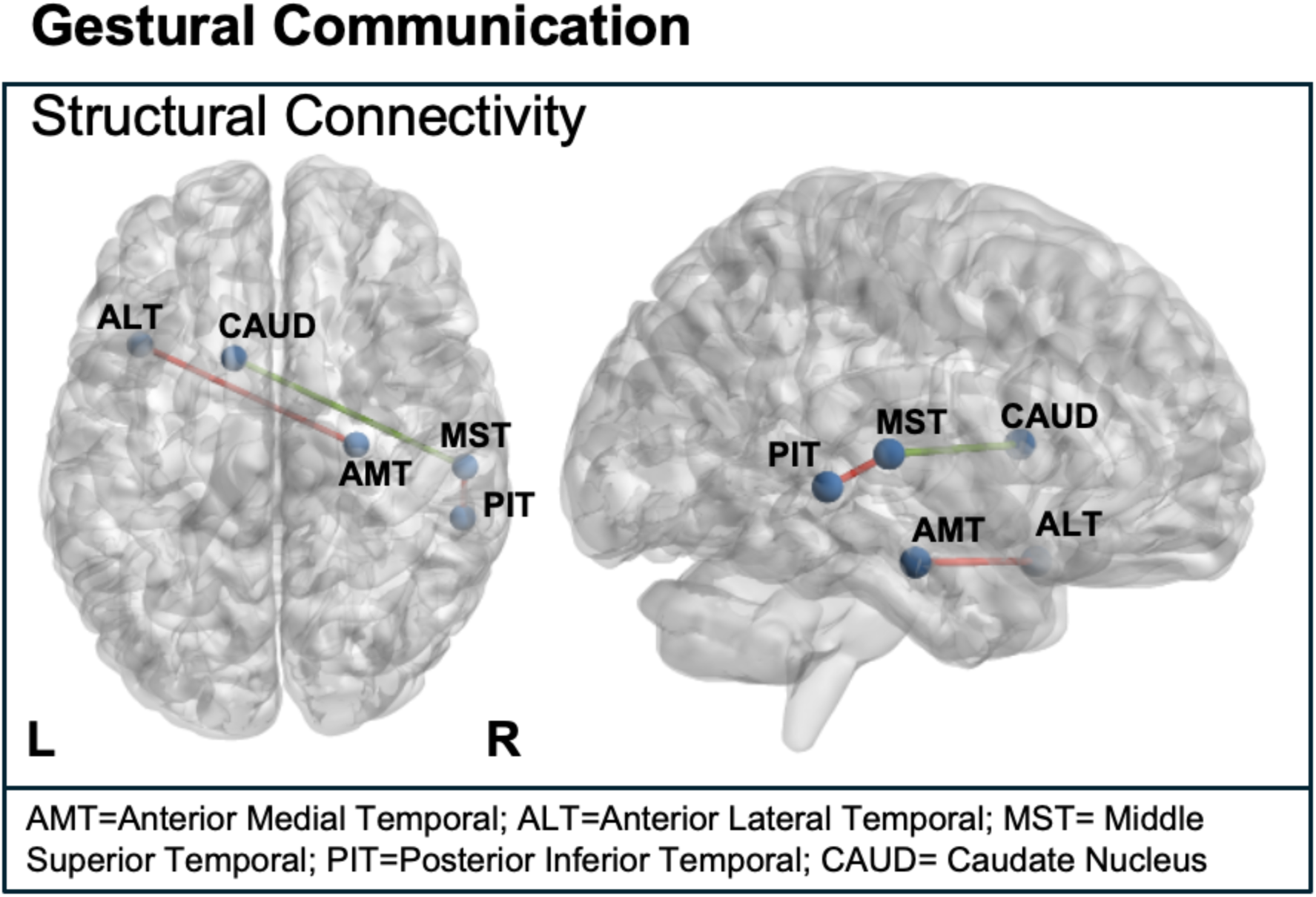
Pairwise structural connections at TEA associated with Gestural Subscale raw scores at age 2. Red = positive associations, Green = negative associations. There were no functional connections that showed significant associations.

**Figure 3.**
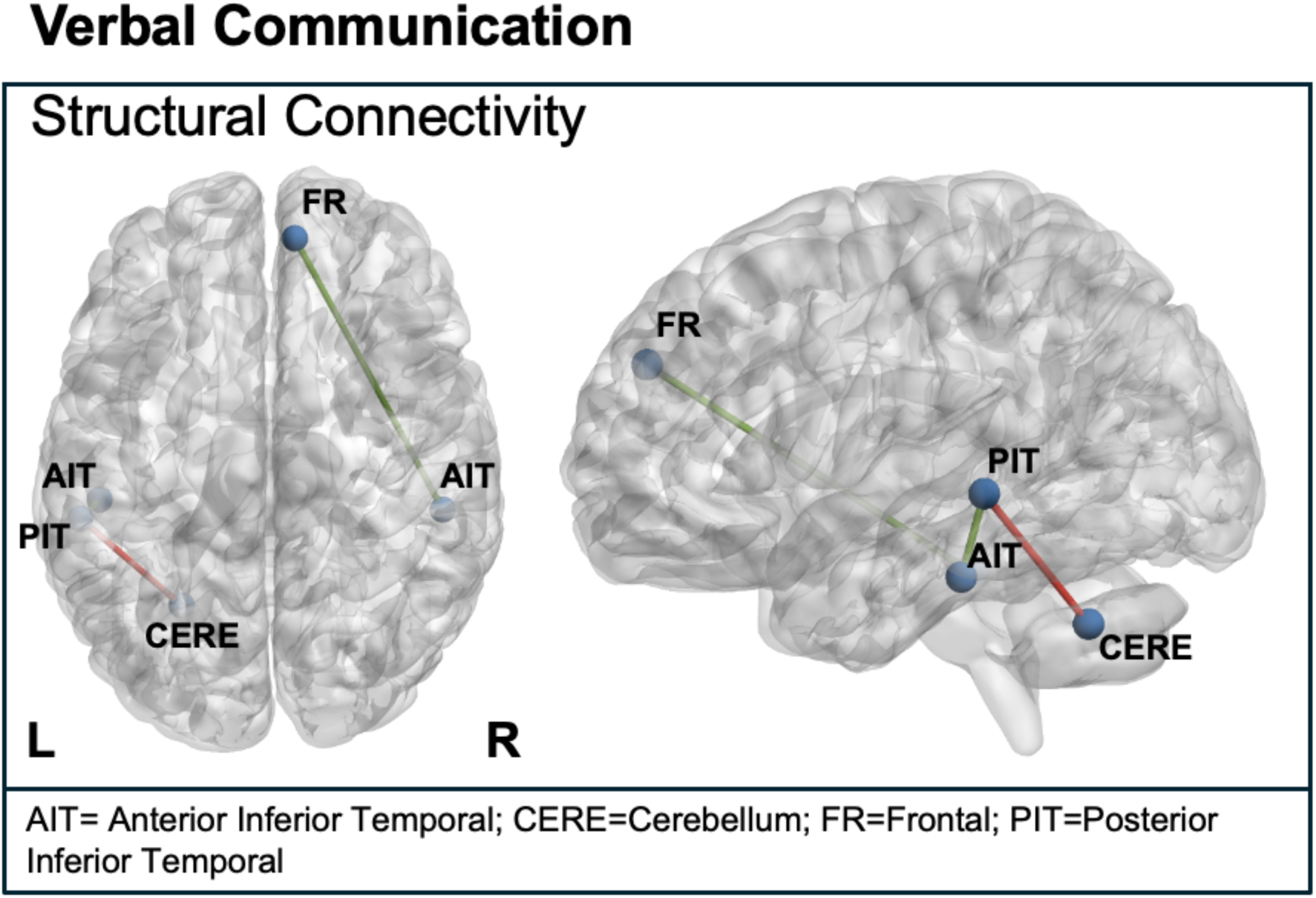
Pairwise structural connections at TEA associated with Verbal Subscale raw scores at age 2. Red = positive associations, Green = negative associations. There were no functional connections that showed significant associations.

**Figure 4.**
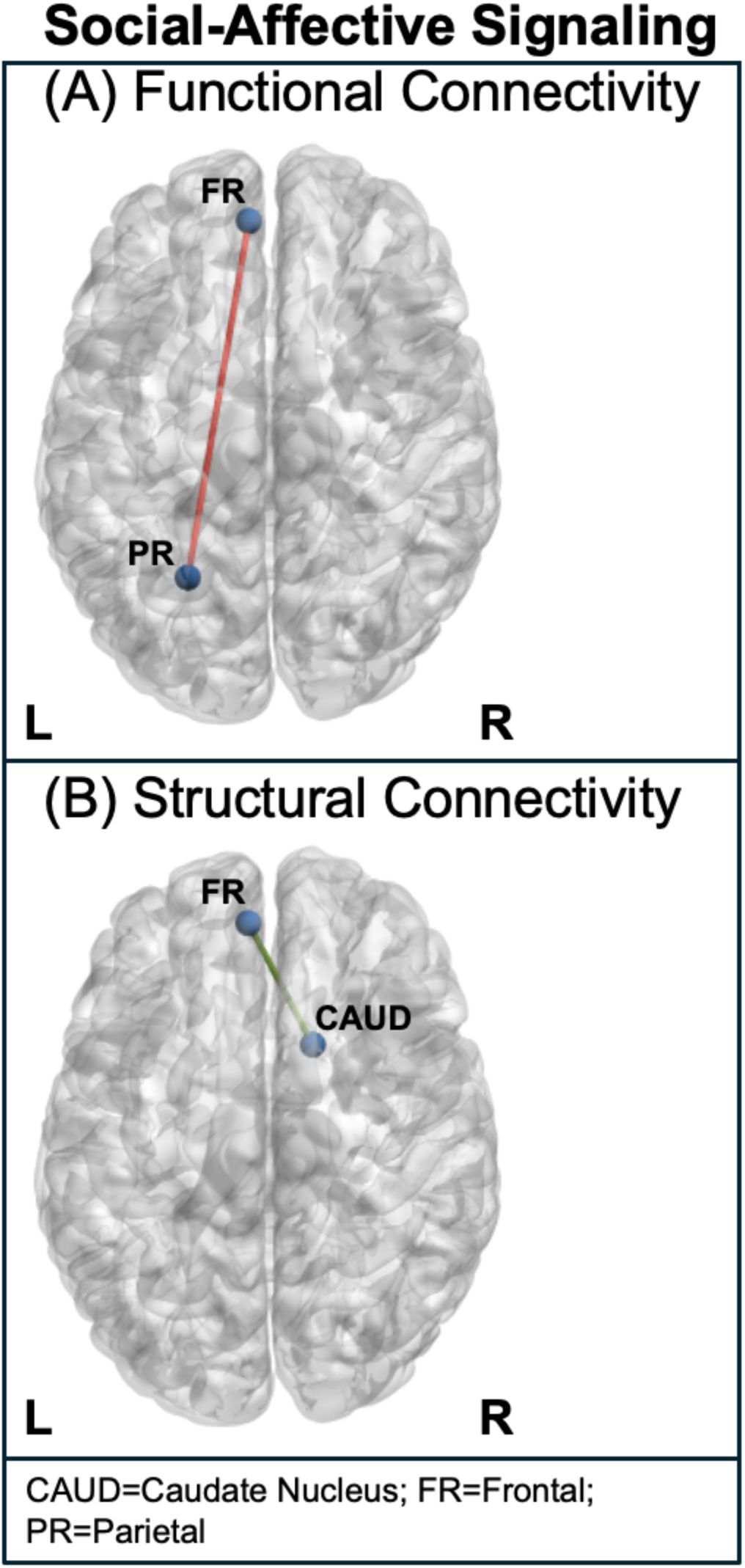
Pairwise functional and structural connections at TEA associated with Social-Affective Subscale raw scores at age 2. Red = positive associations, Green = negative associations

**Figure 5.**
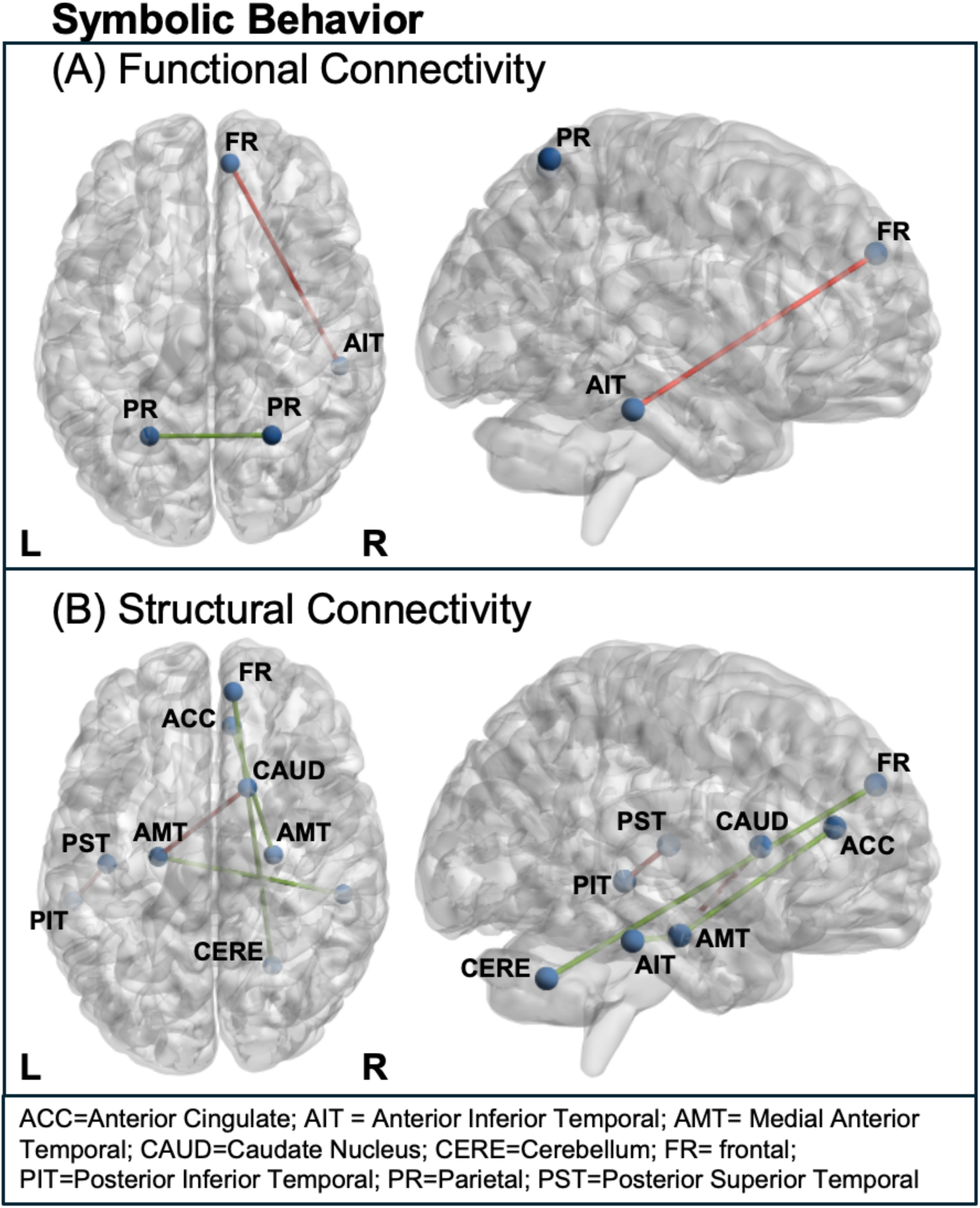
Pairwise functional and structural connections at TEA associated with Symbolic Behavior Subscale raw scores at age 2. Red = positive associations, Green = negative associations

## Data Availability

All data produced in the present study are available upon reasonable request to the authors AFTER peer-reviewed publication of the manuscript.

## Acknowledgements

This work was supported by the National Institutes of Health (R01NS094200, R01NS096037, PI Parikh; R01DC018734, PIs Hunter/Vannest).

